# The impact of extreme weather events on the mental health of vulnerable populations in Africa (The WEMA project): a study protocol

**DOI:** 10.64898/2026.03.19.26348772

**Authors:** Nondumiso Mthiyane, Sithembiso Ndlovu, Agnes Kiragga, Frank Tanser, Aditi Bunker, Vasco Cumbe, Isaias Ramiro, Alberto Muanido, Henry Odero, Evans Omondi, Prasad Liyanage, Elisabeth Lindner, Nafissatou Traoré, Ali Sié, Till Barnighausen, Flavian Otieno, G. Nduku Wambua, Lucy Jilly Akinyi, Sammy Khagayi, Chanelle Mulopo, Frederick Murunga Wekesah, Astrid Treffry-Goatley, Gillian F. Black, Collins Iwuji

**Author notes:** **Corresponding author:** Prof Collins Iwuji, Africa Health Research Institute, KwaZulu-Natal, South Africa.

## Abstract

**Background:** Extreme weather events (EWEs) are increasing in frequency and intensity due to climate change. EWEs negatively affect both physical and mental health, with vulnerable populations disproportionately impacted. Limited data on the specific effects of EWEs on mental health in Africa highlights the need for more research to guide policy and practice. The WEMA study aims to explore the impact of EWEs, in particular storms, cyclones, flooding, and heavy rainfall on common mental disorders (CMDs) in Burkina Faso, Kenya, Mozambique, and South Africa.

**Methods:** This study will employ a transdisciplinary research approach integrating qualitative and quantitative methods to generate contextually grounded and policy-relevant evidence on the mental health impacts of EWEs in sub-Saharan Africa (SSA). We begin with Preferred Reporting Items for Systematic reviews and Meta-Analyses (PRISMA) guided rapid literature review to synthesise existing evidence on the relationship between EWEs and mental health. Secondary analysis of health and demographic surveillance system (HDSS) data across multiple African sites will assess the temporal association between temperature, precipitation, and mental health-related morbidity and mortality, using time series regression with distributed lag non-linear models. In parallel, cross-sectional surveys will estimate the prevalence of CMDs among adults exposed and unexposed to flooding. Logistic regression, accounting for confounders, will be used to estimate odd ratios of the impact of flooding on CMDs. An embedded qualitative study will involve thematic analysis of digital stories produced by community-based co-researchers through participatory workshops, capturing lived experiences of EWEs. Findings from both components will be synthesised and disseminated through knowledge exchange meetings to bridge scientific and experiential insights and inform locally relevant interventions.

**Discussion:** The pool of evidence generated through this transdisciplinary study will be widely shared to draw attention to the impact of EWEs on mental health and to inform relevant policy and practice. Through this work, we aim to advance locally relevant climate adaptation strategies to help reduce health inequalities and support the psychosocial well-being of affected communities.

## BACKGROUND

Climate change is projected to further increase the number and severity of heatwaves, floods and droughts, causing a disproportionate increase in injury, illness and death in resource-limited settings already burdened with wide-ranging health conditions (1). Some population groups (e.g., children, older adults, people with chronic illnesses or mobility challenges, poor or isolated people, and pregnant women) often experience a disproportionate share of the health risks associated with extreme weather events (EWEs) due to the intersecting social vulnerabilities that they face (2–4). The lesser-known, and often overlooked, effects of climate change include the risks and impacts to mental health.

Mental disorders are the second leading cause of years lived with disability (YLD) (5), hence have devastating impacts on individuals, communities, and society. Despite this, there is a poor health system response to the burden of mental disorders with a large gap between the need for and provision of treatment with an estimated 76-85% of people with severe mental disorders receiving no treatment for their disorder in low-and middle-income countries (6). The risks and impacts of climate change on mental health are already rapidly accelerating, resulting in several direct, indirect, and overarching effects that disproportionately affect those who are most vulnerable and marginalised (7). Additionally, studies investigating causal relationships between EWEs on mental health in Africa are rare with only one published study of the impact of cumulative disaster on first onset depression in South Africa (8).

In previous work, we estimated high prevalence of common mental disorders (CMDs) in different population groups including adolescent girls and young women in South Africa and Kenya (9, 10) primary health care clinic attendees in Mozambique (11) and health care professionals during the COVID-19 pandemic in Burkina Faso and four other African countries (12). In a study utilising Health and Demographic Surveillance System (HDSS) data collected over a 16-year period in Burkina Faso, we established significant correlations between specific weather patterns and increased mortality risks. Our analysis revealed that high temperatures combined with low precipitation were linked to a higher risk of malaria-related deaths, particularly in children, within a 14-day window. Additionally, we found that elevated temperatures were associated with an increased risk of death from cardiovascular diseases among adults aged 65 and older, with the highest risk observed within one week following temperature spikes (13). This research underscores the potential impact of weather conditions on public health, highlighting the vulnerability of both younger and older populations to EWEs.

In rural South Africa, we applied a systems-thinking framework to explore the complex social, economic, and demographic pathways linking drought conditions to poor adherence to HIV treatment (14–16). The analysis revealed how environmental stressors, such as drought, influence health behaviours and access to care, emphasising the need for a broader understanding of climate-related challenges in shaping health outcomes. In addition, our systematic review of 105 studies found a significant increase in emergency room visits and hospitalisations for mental health disorders during periods of extreme heat (17). However, it is important to note that none of the studies in the review were conducted in Africa, underscoring the urgent need for further research on the relationship between EWEs and mental health in the African context to inform relevant policies and practices. These findings highlight the interconnected impacts of climate change events on public health, illustrating both the direct and indirect ways in which EWEs exacerbate health challenges across different populations. This underscores the importance of adopting integrated, systems-based strategies to address the multifaceted effects of climate change on health, especially in Africa, where vulnerability may be particularly high.

Most studies on climate change and mental health are quantitative in nature. There is a need to explore the lived experiences of people and communities impacted by climate change, especially those living in communities facing social and structural vulnerabilities. Consequently, in WEMA, we will apply the participatory research methodology of digital storytelling (DST) to generate narrative and visual lived experience data on this topic (18, 19). In participatory research, the agency and autonomy of participants is respected by involving them in multiple steps of the research process (20). This approach can help to align the study with public priorities and community needs (21). For example, WEMA community-based co-researchers (CBCRs) recruited at each site, will be directly involved in multiple phases of the project, including data generation (DST), data analysis, knowledge exchange, intervention design, and public and policy engagement.

The aim of this study is to demonstrate the direct effects of EWEs on CMDs, including depression, anxiety, and post-traumatic stress disorder (PTSD), in Burkina Faso, Kenya, Mozambique, and South Africa.

The objectives of the study are to:

1. Rapidly generate new evidence using existing data on the effects of EWEs on CMDs in vulnerable populations.
2. Investigate the burden and impact of CMDs amongst vulnerable populations exposed to EWEs.
3. Engage community, policy and civil society stakeholders in the interpretation of the evidence generated to brainstorm context-specific interventions.
4. Deliver a targeted public engagement and strategic communication campaign to drive policy change to support mental health and wellbeing in partnership with local leaders, community representatives and advocacy groups.

This study protocol details the methodologies for achieving the first two objectives.

## METHODS

### Study design

This study will use a mixed method approach where both qualitative and quantitative methods of data collection and analysis are integrated, enabling the team to better understand community dynamics and evaluate the impact of EWEs on mental health among vulnerable populations. Work packages (WPs) will be used to establish a standardised research implementation process across all countries for comparative purposes.

#### WP1: Rapidly generate new evidence on the effects of EWEs on CMD

##### WP 1.1 Synthesise evidence on the impact of EWEs on the mental health of vulnerable populations in sub-Saharan Africa

We will conduct a systematic literature review to understand the direct impact of EWEs on the mental health of vulnerable populations in sub-Saharan Africa. Following the PRISMA guidelines for systematic literature reviews, three databases will be searched (PubMed, PsycInfo and Web of Science), with different MeSH terms for “mental health”, “extreme events” and “sub-Saharan Africa”, all separated by the appropriate Boolean operator. Standard systematic review methods will be applied aiming to rapidly identify links and evidence gaps in the current literature on the impact of EWEs on mental health.

##### WP1.2. Investigate the impact of EWEs on CMDs and mental-health-related mortality using secondary data

We will examine the relationship between extreme heat and rainfall and their impact on cause-specific mental health-related deaths and illnesses (ICD10 codes F00-99) across HDSS sites in study countries.

Our analysis will utilise anonymised hospital records and self-reported causes of illness, as well as cause of death determined by verbal autopsies at each HDSS site, to assess the temporal associations between daily and weekly maximum temperatures, total precipitation, and daily deaths attributed to mental health outcomes. Environmental data will be obtained from local weather stations closest to each site and complemented with data from the European Centre for Medium-Range Weather Forecasts reanalysis for the global climate and weather version 5 (ERA-5) (22).

#### WP2: The burden of CMDs and impact of EWEs on mental health

##### WP 2.1. Prevalence of CMDs

We will carry out a cross-sectional prevalence survey that will quantify the symptoms of depression, anxiety and PTSD amongst vulnerable populations exposed to EWEs across all study sites.

##### WP 2.2: Digital storytelling

To generate community-derived data on the impacts of exposure to EWEs on mental health, we will apply the participatory arts-based research methodology of DST (18, 19). Digital stories are short multimedia narratives that combine spoken words, visuals, and text to convey a message or share an experience. Participants create stories at facilitated workshops where they learn skills to describe and illustrate their experiences of a particular challenge that is affecting them. This will be our primary participatory data generation methodology. Digital stories are short audiovisual clips created in workshops by research participants about a particular lived experience (23). DST has been applied in multiple contexts to deepen understanding of the lived experiences of a phenomenon under study (18). For example, while Treffry-Goatley and colleagues harnessed this method to explore the lived experience of HIV drug adherence in a rural South African community (18), Black and colleagues have applied this method to learn more about the impact of natural disasters on vulnerable communities in urban, South African settings (24). Additionally, both researchers have used digital stories as evocative engagement tools to support policy engagement and strategic communication (25).

The DSTs will be created at participatory workshops held in the four study sites and will offer key insight into lived experiences of EWEs. Analysis of the DSTs will be undertaken in 2 stages: 1) participatory analysis by the CBCRs at each site, supported by community engagement teams (CET) based at the local academic institutions 2) in-depth thematic analysis by the research team, building on the findings of the participatory enquiry. The results of the DST analysis and the prevalence surveys will be shared at knowledge exchange meetings between CBCRs and local researchers to facilitate the integration of scientific and experiential knowledge. These meetings will be followed by brainstorming and co-design workshops to allow for participatory reflection and the planning of potential interventions.

### Study settings

The study will be implemented in four sites: Beira, Dondo and Nhamatanda, Sofala, Mozambique; Nouna, Kossi, Burkina Faso; Nairobi and Kisumu, Kenya; and eThekwini and Hlabisa, KwaZulu-Natal (KZN), South Africa. These countries are characterised by high vulnerability to climate change and limited capacity to address its challenges (26–29). While South Africa is comparatively less vulnerable and more prepared, it remains the most unequal nation globally (30) resulting in large proportion of the population being at risk. The study will be implemented by the following research partners:

#### Africa Health Research Institute (AHRI)

AHRI is a Wellcome Trust Africa and Asia Programme in KZN, South Africa. It operates from two campuses: an HDSS and clinical research infrastructure located in rural northern KZN, and a world class laboratory infrastructure located in Durban. The DST will be carried out in Durban, eThekwini, specifically in areas impacted by floods where some of the population has been displaced into temporary shelters and the prevalence survey will be carried out in Northern KwaZulu-Natal.

#### The African Population and Health Research Center (APHRC)

APHRC is a research institution and think tank headquartered in Nairobi, Kenya. In 2002, the Center established the Nairobi Urban Health and Demographic Surveillance System (NUHDSS) in two slum communities within Nairobi, Kenya. In Kenya, the WEMA study will be implemented in Mukuru Kwa Reuben, an informal settlement in Nairobi, as well as in Manyatta, a slum in Kisumu, which will serve as the control/unexposed community but with similar socioeconomic characteristics as Mukuru Kwa Reuben.

Mukuru kwa Reuben is one of the largest informal settlements in Nairobi County, Kenya, and forms part of the wider Mukuru informal settlement cluster (31). Positioned along the Ngong River, it is close to Nairobi’s Central Business District, the country’s major economic and administrative hub. The settlement occupies a low-lying floodplain, a geographic characteristic reflected in the name Mukuru, meaning “valley” in the Kikuyu language. As settlements like Mukuru expand into hazardous flood zones, flood exposure increases (32).

Due to its topographical and infrastructural vulnerabilities, including poor drainage systems, inadequate housing, and limited waste management, Mukuru kwa Reuben is highly susceptible to seasonal flooding during periods of heavy rainfall (32). Such events frequently lead to displacement, property damage, and outbreaks of waterborne diseases such as cholera and typhoid (33). The rapid population growth, driven by its proximity to Nairobi’s primary industrial corridor, establishes Mukuru kwa Reuben as a vital source of affordable housing for low-income earners working in the adjacent industrial zones (34).

#### The Centre de Recherche en Sante de Nouna (CRSN)

CRSN is a Burkina Faso national research institute belonging to the National Institute of Public Health in the Ministry of Health. CRSN has a long and solid experience in the design and implementation of research and evaluation of health interventions and operates an HDSS since 2000. The setting of the prevalence survey will be villages within the catchment area of the Nouna HDSS and whose homes have been flooded due to EWEs, and Individuals impacted by EWEs would either currently be residing in temporary accommodation or makeshift shelters.

#### Comité para a Saúde de Moçambique (CSM)

CSM is a local Mozambican non-governmental organisation, formerly Health Alliance International, that works closely with the Ministry of Health in Mozambique in health system strengthening. CSM, together with the Department of Global Health at the University of Washington, Seattle, has been at the forefront of strengthening mental health in primary health care in Mozambique. The setting for the prevalence survey will be Beira, Dondo and Nhamatanda. Individuals who have been exposed to flooding with or without displacement from their usual residence and residing in temporary shelters, camps, or makeshift housing or another neighbourhood would be offered participation in the study. Unexposed individuals will be recruited from Nhamatanda District.

### Study Participants

In the secondary analysis investigating the impact of EWEs on CMDs and mental-health-related mortality using verbal autopsy and hospital information system data from the HDSS sites contributing data (WP1.2), participants (no age restrictions) with complete observations will be included in the analysis.

To quantify the prevalence of symptoms of depression, anxiety and PTSD amongst vulnerable populations exposed to EWEs (WP2.1), we will conduct a cross-sectional prevalence study of CMDs amongst a sample of individuals in i) KwaZulu-Natal, South Africa; ii) Beira, Dondo and Nhamatanda in Sofala, Mozambique, iii) Nouna in Burkina Faso; and iv) Mukuru Kwa Reuben slum in Nairobi and Manyatta slum in Kisumu, Kenya. Participants will include exposed (flooded homes) and may or may not have experienced displacement and unexposed/not displaced individuals with respect to EWEs. We will include individuals:

- aged 18 years and above,
- resident in the setting identified for recruitment for at least 6 months,
- who have the mental capacity to participate in the study and can give informed consent.

We will exclude individuals if they:

- lack capacity to provide informed consent,
- are at their last trimester of pregnancy or at 3-months post-partum.

### Sampling and Recruitment

For prevalence analysis (WP2.1), recruitment of participants will be tailored to each context but will follow a random sampling strategy. In South Africa, following recent flooding events, all households affected by flooding within the HDSS will be identified. A sampling frame will be constructed from these households, from which one flood-exposed individual will be randomly selected per household. An analogous approach will be used to identify and sample unexposed individuals within the HDSS

In Kenya, for the exposed group, which is Mukuru Kwa Reuben, we will work with the community guides to randomly select villages located along Ngong River that have experienced flooding. We will focus on households or people whose floodwater entered their homes, and they may or may not be displaced. The households or people will be randomly chosen from these villages for the study. We will use a similar method for the unexposed community (Manyatta A) to identify individuals whose homes have not been flooded.

In Mozambique, the exposed/displaced group (Beira City: Praia Nova, Ndunda 2 and Póvoa; Dondo City: Mútua and Mandruzi) will be selected because they are places that are cyclically affected by heavy rainfalls resulting in temporary displacement from their usual residences. In Praia Nova, people live in low-lying areas close to the beach and are very prone to flooding. Those residing in Ndunda 2 live in low-lying areas and cyclically suffer from rainwater flooding. Póvoa is a floodplain with no rainwater collection system, hence prone to flooding. Mútua, located in the Dondo District, is a place where people from Beira and Dondo have been resettled. Mandruzi, is a neighbourhood in the Dondo District with resettled people from Thundani, Mafambisse and Beira City. The unexposed/control group (Nhamatanda District) was chosen because it is a location with sociodemographic characteristics similar to those of people living in the study areas of Beira and Dondo, but without exposure to flooding. These are people living on the edge of national road number 6, in Lamego, Nhamatanda, Nharochonga and Siluvo. We will map houses in the identified areas, where one person from each mapped house will be selected to participate in the interviews. The houses will be mapped starting from the entrance of each neighbourhood to the southernmost end of each neighbourhood. Within each selected house, only one adult will be interviewed. The exposed group will include families in Praia Nova, Ndunda 2, and Póvoa who suffered severe flooding, water infiltration into homes, forcing residents to leave. Families living in the Accommodation Centers at Mútua and Mandruzi are those who have been displaced from their homes in Beira or Dondo due to the floods. The unexposed group will include families living in the Nhamatanda District on the stretch between Lamego, Vila de Nhamatanda, Nharochonga, and Siluvo. These are families with sociodemographic characteristics similar to those living in Beira (Praia Nova, Ndunda 2 and Póvoa) and Dondo (Mútua and Mandruzi) but without exposure to flooding and without having moved from their places of residence in the last 12 months.

In Nouna, we identified 13 villages within the HDSS of Nouna that experienced flooding during the past three years. In the first stage, the sampling frame will be constructed at the household level, using field enumerators who list households affected by flooding in these villages (Household ID and the name of the head of household collected during this process). The HDSS database provides socio-demographic information of each individual (names, ages, ethnicity, religion, education, etc.) residing in the household.

In the second stage, all eligible individuals will be selected and preloaded onto tablets using HDSS data. This will assist data collectors during the household visit to identify eligible adults for selection and ensure the interview is conducted in a household that was affected by flooding. Only one eligible person will be randomly selected per household. A third of participants will be between the ages of 18−24.

Importantly, the selected individual must have been personally exposed to flooding in the selected household. Data collectors will confirm the number of eligible adults and verify exposure before proceeding with selection, informed consent, and data collection.

For the control group, households that were not affected by flooding will be identified within the same areas using the HDSS database. As with the exposed group, a list of eligible household members will be preloaded onto the tablets for each control household. During the household visit, data collectors will randomly select one eligible adult to participate in the interview. The selected individual must not have been affected by flooding in the past three years, meaning they resided in a household that was not flooded during that time. Exposure status will be confirmed before informed consent and data collection. Similar to the exposed group, a third of the participants in the control group will be aged 18−24.

### Sample size

From previous studies, prevalence of depression in study countries ranged from 3.6% to 4.6%, and anxiety from 2.7% to 3.4% based on the global burden of disease study in 2015 (35). Studies in South Africa and Mozambique estimated the prevalence of PTSD to be 2.3% and 4.8% respectively (11, 36). We assumed a higher prevalence of 7.5% among vulnerable unexposed populations. Assuming the impact of EWEs doubles the prevalence to 15%, with 80% power, we anticipate recruiting 278 participants per group (exposed vs. unexposed). However, the nature and impact of EWEs affect countries and their communities differently which we accounted for by assuming an inter-cluster correlation coefficient of 0.01 and a design effect of 1.2, increasing the sample size to 333 participants per group. We estimated a 15% refusal/non-response rate, and a final sample size was 833 individuals (417 in each group, where 138 should be aged 18−24 years old).

For WP2.2, we will be using purposive sampling to identify potential participants for the DST workshops. Ten community members with lived experience of flooding from each site will be selected with the help of the community engagement teams and invited to attend a DST workshop.

### Data collection

In this section, we describe the data extraction/collection and management for the literature review (WP1), secondary data analysis (WP1.2), prevalence survey (WP2.1), and digital storytelling (WP2.2).

#### WP1: Rapidly generate new evidence on the effects of EWEs on CMDs

Two reviewers will independently screen titles and abstracts to exclude studies that do not meet the inclusion criteria. They will also assess the full text of potentially relevant studies, resolving any discrepancies through discussion or with the help of a third reviewer if needed. A standardised data extraction form will be utilised to gather information on study characteristics (such as authors, publication year, study design, and sample size), population details (demographic characteristics and vulnerability factors), specifics of EWEs (type, severity, and duration), and mental health outcomes (types of mental health issues assessed and the measurement tools used).

#### WP1.2. Investigate the impact of EWEs on CMDs and mental-health-related mortality using secondary data

Data extraction will include the following variables

- Social and demographic variables collected from household members in the HDSS sites.
- Anonymised hospital records or self-reported cause of illness.
- Cause of death data (ICD-10 codes F00-99) collected through verbal autopsy or hospital records.
- Daily and weekly maximum temperature, minimum temperature, and total precipitation data.
- Available local meteorological stations and global gridded climate datasets.

#### WP2.1 Quantify the prevalence of symptoms of depression, anxiety and PTSD amongst vulnerable populations exposed to EWEs

Data will be collected through structured questionnaires administered by trained clinical research assistants using Android tablets. To ensure effective data collection, the questionnaires will be translated into the local language. All collected data will be stored on Research Electronic Data Capture (REDCap) web application, with access restricted to authorised study personnel only. Personal identifying information will not be included in the study databases, and all records will be securely stored in password-protected files.

#### Exposure variables

The exposed group will consist of individuals exposed to EWEs such as floods, cyclones, and torrential rains. They may or may not have been displaced as a result of the event.

The unexposed group will include individuals who were not directly impacted by flooding but have similar characteristics to individuals in the exposed group.

#### Confounders

We will collect data on potential confounders including age, sex, pre-existing illness, socio-economic status, ethnicity, marital status, education, employment, safety and security, household composition, food security measured using the U.S. household food security module, alcohol consumption measured using the World Health Organization (WHO) alcohol use disorder identification tool (AUDIT-C) (37), recreational drug use measured using the drug abuse screening test, self-reported HIV status, quality of life using EuroQoL scale (38), healthcare utilisation, adverse childhood experience questionnaire (39), self-efficacy measured using the general self-efficacy scale (40), and social support measured using the multidimensional scale of perceived social support (41).

#### Outcomes

For mental health outcomes, the following clinical screening tools will be used; the Patient Health Questionnaire (PHQ-9) for depression, Generalised Anxiety Disorder scale (GAD-7) for anxiety (42, 43) and PTSD checklist of DSM-5 (PCL-5) for PTSD (44, 45). Cut-off scores were ≥10 for PHQ-9/GAD-7 and ≥31-33 for PCL-5 (44, 45). Severity will be assigned based on validated cut-off scores. The PHQ-9 and GAD-7 have been shown to be effective screening tools for depression and anxiety in African settings, but there is a lack of validated tools for PTSD (42, 43). We recommend using the PCL-5, which has been validated in Zimbabwe and demonstrated strong psychometric properties, with a cut-off score of 33 showing reliable performance (46, 47).

##### WP2.2: Digital storytelling

Data will be generated through digital storytelling workshops attended by 10 CBCRs at each site. The workshops will be led by two experienced digital storytelling facilitators with the support of the local CETs and at least one mental health professional (trained Counsellor). In WEMA, each CBCR will produce their own digital story, a video of 2−6 minutes duration, conveying a personalised lived experience of how their mental health and wellbeing has been impacted by an extreme weather event they have experienced in the past 2−3 years. Participants will be asked to each create a story in response to the following prompt:

*Tell us a true story about a time in the recent past (last 2−3 years) when you experienced an extreme weather event that affected your mental health. We would like to understand how the way you were thinking or feeling influenced your ability to get through that event. Is there anything that stands out in your memory such as something that happened or something that you or someone else did that made it more difficult or made it easier for you to bear the weather event?*

i. The stories will be generated during a 5-day workshop which will be held in all four sites. Day 1 involves topic orientation and story identification; Day 2 focuses on story development; Day 3 is dedicated to completing the story and creating initial images; Day 4 involves artwork creation; and Day 5 is for finalising and digitising the artwork.

Outputs: Transcribed workshop dialogue, approximately 10 DSTs per site, the DST narratives and accompanying visual imagery, academic manuscripts.

Outputs: Causal Loop Diagrams (CLDs), recorded discussion.

### Analysis

#### Data synthesis (literature review)

##### WP1: Rapidly generate new evidence on the effects of EWEs on CMDs

This will involve a narrative synthesis of data extracted from the articles to summarise the findings. Data synthesis will involve grouping the studies by type of common mental disorders and EWEs, identifying common mental health outcomes across studies, and highlighting key themes and patterns in the data and elucidating pathways linking EWEs to mental health disorders. The quality of the included studies will be assessed using existing tools such as the Newcastle-Ottawa Scale for observational studies (48) and the Cochrane Risk of Bias Tool for randomised controlled trials (49). The review will be reported following the PRISMA (Preferred Reporting Items for Systematic reviews and Meta-Analyses) guidelines, ensuring transparency and reproducibility.

#### Statistical analysis

##### WP1.2 Investigate the impact of EWEs on CMD and mental-health-related mortality using secondary data

We will analyse anonymised daily hospitalisation and cause of death data from each HDSS site to evaluate the temporal associations between daily and weekly maximum temperatures, total precipitation, and mental health-related deaths. Time series regression analysis will be conducted using distributed lag non-linear models for mental health outcomes, generating effect estimates that consider lagged impacts of up to 30 days. We will compute exposure-response relationships for precipitation and maximum temperature concerning mental health for each HDSS site and then combine these estimates using meta-regression techniques to identify the best linear unbiased estimates.

##### WP2.1 To quantify the prevalence of symptoms of depression, anxiety and PTSD amongst vulnerable populations exposed to EWEs

We will quantify the prevalence of depression, anxiety, and PTSD symptoms among populations exposed to EWEs, expressed as a percentage of the total within each exposure category. We will also estimate the prevalence of symptoms of any CMD. Logistic regression models, adjusting for potential confounders, will be used to measure the association between EWEs and mental health outcome, and quantified using odds ratio. All pre-identified confounders will be included in the models regardless of their statistical significance. For sensitivity analysis, we will use linear regression where outcomes will be included as continuous variables. After calculating estimates for each country individually, we will use meta-regression techniques to combine these estimates.

##### WP2.2 Digital storytelling

Multiple layers of data will be produced through the DST workshops, including the DSTs, transcribed narratives, visual data, and transcribed workshop discussions. The analysis approach will be based on the principles of thematic analysis (TA), where descriptions of emerging themes are produced through a process of coding and systematising data (50–52). In our application of participatory TA, we will draw on the six steps to TA outlined by Braun and Clarke, including: i) familiarisation with data, ii) generating initial codes, iii) identifying themes that reflect collections of codes, iv) reviewing data to understand and explain the meaning and dynamics of themes, v) maintaining rigor through inter-coder agreement, vi) producing the final report (51). The first four steps will be carried out by the CBCRs at participatory workshops held at each site under the guidance of the CET. The CBCRs will collectively analyse their collection of stories and will engage in an accessible process of data coding to generate themes. This will ensure that the CBCRs interpret and unpack their own stories and make meaning of them in ways that they wish them to be understood by others. The last two steps will be carried out by the research team. The initial themes produced by the CBCRs will inform further layers of thematic analysis, synthesising the data from the total pool of 40 digital stories generated across all four sites. For this analysis, the transcribed DST data will be uploaded to ATLAS.ti software for qualitative data analysis.

## DISCUSSION

This transdisciplinary study will build critical evidence related to the impact of climate change on mental health in sub-Saharan Africa. Our results will be generated through rigorous mixed methods research approaches and will make a significant contribution to the development of mental health interventions that aim to support populations that are vulnerable to the impacts of EWEs. In addition, the study will provide valuable insights into how mental health services can be better prepared to address the growing challenges posed by climate change.

Furthermore, the results will inform efforts to mitigate the harmful effects of EWEs on mental health conditions that are sensitive to climate variations, such as anxiety, depression, and PTSD. In addition to these practical applications, the study will also identify critical gaps in existing data systems, highlighting areas that need improvement to enhance the effectiveness of future research on climate and mental health. This will help shape the development of more robust, evidence-based strategies to support mental health in the context of a changing climate.

Most CMDs in adults have their onset during childhood. However, in WP 2, we have made the decision to exclude participants under the age of 18 from our study. This choice is driven by our intention to protect these younger individuals from discussing potentially distressing topics, such as flooding, which could inadvertently retraumatise them or worsen any existing mental health conditions. Furthermore, our team lacks the presence of a child psychologist, who would typically be needed to offer appropriate support and counselling to any children who might experience emotional distress during the study. By excluding minors, we aim to prioritise their well-being and ensure that participants have the necessary resources to cope with sensitive subjects. While this decision safeguards the welfare of younger individuals, it may limit the generalisability of our findings, particularly in relation to the impact of EWEs on mental health across the lifespan. The exclusion of children means that we will not capture how these events affect younger populations, which could offer valuable insights into the early onset of mental health issues linked to environmental stressors. This gap may influence the overall conclusions about the long-term mental health outcomes of weather-related trauma. We have developed a distress protocol, including access to a psychologist to address the issue of re-traumatisation in adults.

A notable strength of the prevalence survey is the inclusion of a control group which allows us to measure background prevalence of CMDs in the studied population. This approach allows for the measurement of the relative contribution of flooding to the development of CMDs, which is lacking in previous studies in SSA to date (53).

## Data Availability

All data produced in the present study are available upon reasonable request to the authors.

## List of abbreviations

AUDIT-C: Alcohol Use Disorder Identification Tool
CBCRs: Community-based co-researchers
CET: Community engagement team
CHW: Community health worker
CLD: Causal loop diagram
CMD: Common mental disorder
DST: Digital storytelling
EoI: Expression of interest
EWE: Extreme weather event
GAD: Generalised anxiety disorder
HDSS: Health and demographic surveillance system
KZN: KwaZulu-Natal
NUHDSS: Nairobi Urban Health and Demographic Surveillance System
PCL: Post-traumatic stress disorder checklist
PHQ: Patient Health Questionnaire
PTSD: Post-traumatic stress disorder
TA: Thematic analysis
WHO: World Health Organization
WP: Work package
YLD: Years lived with disability
SSA: sub-Saharan Africa
PRISMA: Preferred Reporting Items for Systematic reviews and Meta-Analyses

## DECLARATIONS

### Ethics approval and consent to participate

This protocol has been reviewed and approved by the University of KwaZulu-Natal Biomedical Research Ethics Committee (BREC) (reference number: BREC/00007752/2024) and gatekeeper permission was obtained from the Department of Social Development in KZN to undertake the study, Comité d’ Ethique pour la Recherche en Santé (CERS) for Burkina Faso (reference number: 2024-12-376), Amref Ethics and Scientific Review Committee (ESRC) for Kenya (reference number: ESRC P1792/2024), and Comité Nacional de Bioética para Saúde’s (CNBS) for Mozambique (reference number: 309/CNBS/25). Participants will be adequately informed about the aims, objectives and modalities of data collection, processing, storage, risks and benefits, and responsibilities pertaining to the WEMA study. They will be provided will the opportunity to ask questions for clarity, and to give their consent clearly and unequivocally before participating in the study. They will also be informed that they can withdraw from participating without any fear of repercussions, and their data can be withdrawn at any time before it is published as journal articles.

### Consent for publication

Participants will be asked to provide consent for their data to be published.

### Authors’ contributions

Conceptualisation: CIW; Funding acquisition: CIW, FT, AB, IR, VC, ATG, GB, AK, AS; Writing-Original draft: NM, SN, CIW; Writing-Review and Editing: all authors. All authors agreed to the final version of the manuscript.

## Acknowledgements

The authors thank the community members whose generosity allow the proposed research to be undertaken by engaging with research teams across the different contexts. This work will not be possible without the support of government agencies and community structures that facilitate access to research communities.

## References

1. Ebi KL, Hess JJ. Health Risks Due To Climate Change: Inequity In Causes And Consequences. Health Aff (Millwood). 2020;39(12):2056–62.

2. Munro A, Kovats RS, Rubin GJ, Waite TD, Bone A, Armstrong B, et al. Effect of evacuation and displacement on the association between flooding and mental health outcomes: a cross-sectional analysis of UK survey data. The lancet planetary health. 2017;1(4):e134–e41.

3. Escobar Carias MS, Johnston DW, Knott R, Sweeney R. Flood disasters and health among the urban poor. Health Economics. 2022;31(9):2072–89.

4. Lundgren M, Strandh V. Navigating a double burden–Floods and social vulnerability in local communities in rural Mozambique. International Journal of Disaster Risk Reduction. 2022;77:103023.

5. GBD 2019 Mental Disorders Collaborators. Global, regional, and national burden of 12 mental disorders in 204 countries and territories, 1990-2019: a systematic analysis for the Global Burden of Disease Study 2019. Lancet Psychiatry. 2022;9(2):137–50.

6. WHO. Comprehensive mental health action plan 2013–2030. Geneva: World Health Organization.; 2021.

7. Hayes K, Blashki G, Wiseman J, Burke S, Reifels L. Climate change and mental health: risks, impacts and priority actions. Int J Ment Health Syst. 2018;12:28.

8. Tomita A, Ncama BP, Moodley Y, Davids R, Burns JK, Mabhaudhi T, et al. Community disaster exposure and first onset of depression: A panel analysis of nationally representative South African data, 2008-2017. PLOS Clim. 2022;1(4):0000024.

9. Erskine HE, Blondell SJ, Enright ME, Shadid J, Wado YD, Wekesah FM, et al. Measuring the Prevalence of Mental Disorders in Adolescents in Kenya, Indonesia, and Vietnam: Study Protocol for the National Adolescent Mental Health Surveys. J Adolesc Health. 2023;72(1s):S71–s8.

10. Mthiyane N, Harling G, Chimbindi N, Baisley K, Seeley J, Dreyer J, et al. Common mental disorders and HIV status in the context of DREAMS among adolescent girls and young women in rural KwaZulu-Natal, South Africa. BMC public health. 2021;21(1):478.

11. Muanido A, Cumbe V, Manaca N, Hicks L, Fabian KE, Wagenaar BH. Prevalence and associated factors of common mental disorders in primary care settings in Sofala Province, Mozambique. BJPsych Open. 2023;9(1):e12.

12. Assefa N, Abdullahi YY, Hemler EC, Lankoande B, Wang D, Madzorera I, et al. Continued disruptions in health care services and mental health among health care providers during the COVID-19 pandemic in five sub-Saharan African countries. J Glob Health. 2022;12:05046.

13. Bunker A, Sewe MO, Sié A, Rocklöv J, Sauerborn R. Excess burden of non-communicable disease years of life lost from heat in rural Burkina Faso: a time series analysis of the years 2000-2010. BMJ Open. 2017;7(11):e018068.

14. Orievulu K, Ayeb-Karlsson S, Ngwenya N, Ngema S, McGregor H, Adeagbo O, et al. Economic, social and demographic impacts of drought on treatment adherence among people living with HIV in rural South Africa: A qualitative analysis. Clim Risk Manag. 2022;36:100423.

15. Iwuji CC, Baisley K, Maoyi ML, Orievulu K, Mazibuko L, Ayeb-Karlsson S, et al. The Impact of Drought on HIV Care in Rural South Africa: An Interrupted Time Series Analysis. Ecohealth. 2023;20(2):178–93.

16. Orievulu KS, Ayeb-Karlsson S, Ngema S, Baisley K, Tanser F, Ngwenya N, et al. Exploring linkages between drought and HIV treatment adherence in Africa: a systematic review. The Lancet Planetary Health. 2022;6(4):e359–e70.

17. Corvetto JF, Helou AY, Dambach P, Müller T, Sauerborn R. A Systematic Literature Review of the Impact of Climate Change on the Global Demand for Psychiatric Services. Int J Environ Res Public Health. 2023;20(2).

18. Treffry-Goatley A, Lessells RJ, Moletsane R, de Oliveira T, Gaede B. Community engagement with HIV drug adherence in rural South Africa: a transdisciplinary approach. Medical humanities. 2018;44(4):239–46.

19. Mpofu-Mketwa TJ, Abrams A, Black GF. Reflections on measuring the soundness of the digital storytelling method applied to three Cape Flats vulnerable communities affected by drought, fire and flooding in Cape Town. Social Sciences & Humanities Open. 2023;7(1):100407.

20. Minkler M, Wallerstein N, editors. Community-Based Participatory Research for Health: From Process to Outcomes. Hoboken, NJ: John Wiley & Sons; 2011.

21. Vaughn LM, Jacquez F. Participatory research methods–choice points in the research process. Journal of participatory research methods. 2020;1(1).

22. Hersbach H, Bell B, Berrisford P, Hirahara S, Horányi A, Muñoz-Sabater J, et al. The ERA5 global reanalysis. Quarterly journal of the royal meteorological society. 2020;146(730):1999–2049.

23. Lambert J. Digital storytelling cookbook. Berkeley, CA: Digital Diner Press; 2010.

24. Black GF, Petersen L, Mpofu-Mketwa TJ, Dick L, Wilson A, Ncube S, et al. Participatory visual methods and the mobilization of community knowledge: working towards community-derived disaster risk management in the context of advancing climate change. Journal of Participatory Research Methods. 2025;6(2):121–52.

25. Lewin T, editor. Digital storytelling: Participatory Learning and Action; 2011.

26. GAIN. Mozambique n.d [Available from: https://gain-new.crc.nd.edu/country/mozambique.

27. GAIN. South Africa n.d [Available from: https://gain-new.crc.nd.edu/country/south-africa.

28. GAIN. Kenya n.d [Available from: https://gain-new.crc.nd.edu/country/kenya.

29. GAIN. Burkina Faso n.d [Available from: https://gain-new.crc.nd.edu/country/burkina-faso.

30. Sulla V, Zikhali P, Cuevas PF. Inequality in Southern Africa: An Assessment of the Southern African Customs Union. Washington, DC: World Bank Group; 2022.

31. Corburn J, Agoe V, Ruiz Asari M, Ortiz J, Patterson R, Ngau P, et al. Situational analysis of Mukuru Kwa Njenga, Kwa Reuben & Viwandani. Technical paper. 2017.

32. Juma B, Olang LO, Hassan MA, Chasia S, Mulligan J, Shiundu PM. Flooding in the urban fringes: Analysis of flood inundation and hazard levels within the informal settlement of Kibera in Nairobi, Kenya. Physics and Chemistry of the Earth, Parts A/B/C. 2023;132:103499.

33. Okaka FO, Odhiambo BD. Households’ perception of flood risk and health impact of exposure to flooding in flood-prone informal settlements in the coastal city of Mombasa. International Journal of Climate Change Strategies and Management. 2019;11(4):592–606.

34. Scott AA, Misiani H, Okoth J, Jordan A, Gohlke J, Ouma G, et al. Temperature and heat in informal settlements in Nairobi. PloS one. 2017;12(11):e0187300.

35. GBD 2015 Disease and Injury Incidence and Prevalence Collaborators ao. Global, regional, and national incidence, prevalence, and years lived with disability for 310 diseases and injuries, 1990–2015: a systematic analysis for the Global Burden of Disease Study 2015. The Lancet. 2015;388:10053.

36. Herman AA, Stein DJ, Seedat S, Heeringa SG, Moomal H, Williams DR. The South African Stress and Health (SASH) study: 12-month and lifetime prevalence of common mental disorders. South African medical journal. 2009;99(5).

37. Saunders JB, Aasland OG, Babor TF, De la Fuente JR, Grant M. Development of the alcohol use disorders identification test (AUDIT): WHO collaborative project on early detection of persons with harmful alcohol consumption-II. Addiction. 1993;88(6):791–804.

38. Rabin R, Charro Fd. EQ-SD: a measure of health status from the EuroQol Group. Annals of medicine. 2001;33(5):337–43.

39. Felitti VJ, Anda RF, Nordenberg D, Williamson DF, Spitz AM, Edwards V, et al. Relationship of childhood abuse and household dysfunction to many of the leading causes of death in adults: The Adverse Childhood Experiences (ACE) Study. American journal of preventive medicine. 1998;14(4):245–58.

40. Schwarzer R, Jerusalem M. Generalized self-efficacy scale. J Weinman, S Wright, & M Johnston, Measures in health psychology: A user’s portfolio Causal and control beliefs. 1995;35(37):82–003.

41. Zimet GD, Dahlem NW, Zimet SG, Farley GK. The multidimensional scale of perceived social support. Journal of personality assessment. 1988;52(1):30–41.

42. Bhana A, Rathod SD, Selohilwe O, Kathree T, Petersen I. The validity of the Patient Health Questionnaire for screening depression in chronic care patients in primary health care in South Africa. BMC psychiatry. 2015;15:1–9.

43. Kroenke K, Spitzer RL, Williams JB, Löwe B. The patient health questionnaire somatic, anxiety, and depressive symptom scales: a systematic review. General hospital psychiatry. 2010;32(4):345–59.

44. Blevins CA, Weathers FW, Davis MT, Witte TK, Domino JL. The posttraumatic stress disorder checklist for DSM-5 (PCL-5): Development and initial psychometric evaluation. Journal of traumatic stress. 2015;28(6):489–98.

45. Bovin MJ, Marx BP, Weathers FW, Gallagher MW, Rodriguez P, Schnurr PP, et al. Psychometric properties of the PTSD checklist for diagnostic and statistical manual of mental disorders–fifth edition (PCL-5) in veterans. Psychological assessment. 2016;28(11):1379.

46. Verhey R, Chibanda D, Gibson L, Brakarsh J, Seedat S. Validation of the posttraumatic stress disorder checklist–5 (PCL-5) in a primary care population with high HIV prevalence in Zimbabwe. BMC psychiatry. 2018;18:1–8.

47. Verhey R, Brakarsh J, Gibson L, Shea S, Chibanda D, Seedat S. Potential resilience to post-traumatic stress disorder and common mental disorders among lay health workers working on the friendship bench programme in Zimbabwe. Journal of Health Care for the Poor and Underserved. 2021;32(3):1604–18.

48. Wells GA, Shea B, O’Connell D, Peterson J, Welch V, Losos M, et al. The Newcastle-Ottawa Scale (NOS) for assessing the quality of nonrandomised studies in meta-analyses. 2000.

49. Higgins JP, Altman DG, Gøtzsche PC, Jüni P, Moher D, Oxman AD, et al. The Cochrane Collaboration’s tool for assessing risk of bias in randomised trials. BMJ. 2011;343:d5928.

50. Boyatzis RE. Transforming qualitative information: Thematic analysis and code development: Sage; 1998.

51. Braun V, Clarke V. Using thematic analysis in psychology. Qualitative research in psychology. 2006;3(2):77–101.

52. Guest G, MacQueen KM, Namey EE. Applied thematic analysis: sage publications; 2011.

53. Deglon M, Dalvie MA, Abrams A. The impact of extreme weather events on mental health in Africa: A scoping review of the evidence. Science of the total environment. 2023;881:163420.

